# Anterior-superior hypothalamic enlargement as specific marker in episodic migraine: converging evidence from an independent discovery-replication design

**DOI:** 10.64898/2026.06.12.26355501

**Authors:** Dan Liu, Shengkun Peng, Longlin Yin, Xueyang Wen, Bin Huang, Keith Maurice Kendrick, Benjamin Becker, Dezhong Yao, Stefania Ferraro

**Author notes:** Corresponding authors: Stefania Ferraro, Dezhong Yao.

## Abstract

**Background:** Growing evidence implicates the hypothalamus as a key structure in migraine pathophysiology; however, our understanding of its precise role and of the specific nuclei involved remains limited. We combined MRI data from our laboratory with publicly available MRI datasets from OpenNeuro to examine hypothalamic subunit volumes in episodic migraine and assess the specificity of these alterations relative to chronic pain conditions.

**Methods:** Structural MRI combined with an automated atlas-based segmentation algorithm and a discovery-replication design was employed to investigate cross-sectional volumetric differences across 5 bilateral hypothalamic subunits in two independent migraine cohorts: DS1-MIG (DS1-MIG-base, n = 111 patients, n = 35 controls) and DS2-MIG (n = 27 patients, n = 31 controls). The adjusted volumes were compared between groups using MANOVA as an omnibus test, followed by Welch t-tests to test univariate follow-up. Longitudinal volumetric changes were additionally assessed in DS1-MIG participants with available follow-up scans using linear mixed models. To assess the specificity of findings to migraine, the same pipeline was applied to two chronic pain datasets, one including patients with fibromyalgia (DS-FM, n = 33 patients, n = 33 controls) and the other including patients with trigeminal neuralgia (n = 119 patients, n = 55 controls).

**Results:** MANOVA revealed significant multivariate group differences in the discovery and replication migraine cohorts (DS1-MIG-base: *p* = .006; DS2-MIG: *p* = .008). Follow-up univariate analyses identified a consistent enlargement of the left anterior-superior subunit across both cohorts (*p*_FDR_ = .023 in DS1-MIG-base and *p*_FDR_ = .046 in DS2-MIG), representing the only cross-cohort replication finding. Beyond this shared signature, DS2-MIG exhibited additional significant enlargements of the right anterior-inferior and right tubular-inferior subunits. Longitudinal analyses in DS1-MIG showed that hypothalamic subunit volumes remained broadly stable over time within both migraine patients and control participants. No significant volumetric alterations were detected in the fibromyalgia or trigeminal neuralgia cohorts, either in multivariate or univariate analyses, underscoring migraine-specific findings.

**Conclusions:** These findings provide evidence for subunit-specific hypothalamic structural alterations in migraine localized in the left anterior hypothalamic subunit. The stability of these differences over time and their absence in other chronic pain conditions suggest a migraine-specific structural organisation of hypothalamic circuitry.

## 1. Introduction

Migraine is a complex neurovascular disorder affecting over one billion people worldwide (Vos et al., 2017; Ashina et al., 2021), whose pathophysiological mechanisms remain poorly understood (Christensen et al., 2025; Karsan and Goadsby, 2025). Current evidence points to the involvement of both the peripheral trigeminovascular system and the central nervous system, with a potential prominent role of the hypothalamus (Ashina et al., 2021). Anatomically, the hypothalamus is a heterogeneous diencephalic region composed of multiple specialized nuclei that collectively support homeostatic, neuroendocrine, autonomic, circadian, and motivated behavioral regulation (Saper and Lowell, 2014).

Crucially, characteristic features of migraine, such as rhythmic recurrence of attacks, the presence of prodromal manifestations (albeit their existence remains debated, see Christensen et al. (2025)), such as yawning, fatigue, and mood alterations, autonomic symptoms like nausea and vomiting, and sensitivity to specific triggers, such as hormonal fluctuations and stress, point to a major involvement of the hypothalamus (May and Burstein, 2019). The earliest direct evidence for a specific involvement of this structure in migraine pathophysiology was provided by Denuelle et al. (2007), who demonstrated sustained hypothalamic activation during spontaneous migraine attacks, together with midbrain and pontine regions that persisted beyond the headache phase. Subsequent studies have consistently reported hypothalamic activation during the premonitory phase and at attack initiation (Maniyar et al., 2014; Schulte and May, 2016; van Oosterhout et al., 2021), suggesting that hypothalamic dysfunction may act as a key driver of migraine onset (May and Burstein, 2019). Numerous additional findings further corroborate this notion. Resting-state regional cerebral blood flow was shown to decrease in the lateral hypothalamus immediately prior to a migraine attack, accompanied by reduced functional connectivity between the hypothalamus and key nodes of the pain-processing network (Meylakh et al., 2020). In a longitudinal study involving multiple scanning sessions, Stankewitz et al. (2021) reported cyclic changes in brain perfusion within the insula and nucleus accumbens, with a peak during the headache phase. In parallel, hypothalamic connectivity to these regions progressively increased across the interictal period toward the attack, followed by a sudden drop during the headache phase, which was interpreted as reflecting a loss of hypothalamic control over limbic system activity. More recently, in a large cohort of episodic migraine patients, Messina et al. (2022) showed that the interictal state is characterized by a widespread reduction in hypothalamic functional connectivity with limbic, sensory, and prefrontal regions, suggesting a baseline dysregulation of hypothalamic networks. Notably, they also showed that increased attack frequency correlated with reduced functional connectivity between the hypothalamus and the lingual gyrus. These findings are paralleled by evidence from preclinical studies identifying molecular pathways involving hypothalamus-derived neuropeptides and neurotransmitters, such as orexins, neuropeptide Y, PACAP, oxytocin, vasopressin, and dopamine, that are implicated in migraine mechanisms (for a recent review, see Salinas-Abarca et al. (2025)).

However, while clinical, functional neuroimaging, and preclinical evidence support a central role of the hypothalamus in migraine pathophysiology, consensus regarding the specific involvement of distinct hypothalamic nuclei and subregions remains elusive. In this context, high-resolution structural MRI provides a non-invasive in vivo approach to investigate whether migraine is associated with hypothalamic macrostructural alterations. An early voxel-based morphometry study reported region-specific alterations with respect to control subjects, showing reduced posterior hypothalamic volume in episodic migraine without aura (n = 18) scanned at least 3 days after the last migraine attack, while reduced anterior hypothalamic volume in chronic migraine (n = 16) (Chen et al., 2019). Two very recent investigations (Giardina et al., 2025; Xiong et al., 2025) employed the same FreeSurfer-based algorithm (Billot et al., 2020), a fully automated segmentation method that has been widely used to investigate hypothalamic structure in both neurological and neuropsychiatric disorders and healthy populations (Abuaf et al., 2022; Luo et al., 2024a; Xie et al., 2023; Hu et al., 2025; Ferraro et al., 2024; Rasmussen et al., 2024), to extract the volumes of 5 bilateral hypothalamic subunits: anterior-superior, anterior-inferior, posterior, tubular-inferior, and tubular-superior.

Xiong et al. (2025) investigated a cohort of migraine patients (chronic, n = 53; episodic, n = 23) irrespective of ictal stage and aura status and reported that, compared with healthy controls, patients with chronic migraine exhibited significant volume reductions in the right superior tubular subunit and in inferior hypothalamic regions (specifically, the left inferior tubular and right anterior inferior subregions). In contrast, patients with episodic migraine did not show statistically significant differences relative to controls, although descriptive analyses suggested a gradual decrease in hypothalamic volumes from controls to episodic and chronic migraine patients within the inferior tubular subunit. The authors speculated that the limited sample size of the episodic migraine group may have resulted in insufficient statistical power to detect significant differences. Notably, the episodic migraine group experienced a mean of ≈ 6 headache days per month. The same study also performed a Mendelian randomization study suggesting that a smaller inferior tubular subregion volume is linked to increased migraine risk (Xiong et al., 2025).

Critically, Giardina et al. (2025) found no significant volumetric differences between a group of 30 episodic migraine patients without aura in the interictal state with respect to a well-matched control group. In this case, migraine patients experienced a mean of ≈ 5 − 6 headache attacks per month.

Altogether, current volumetric findings suggest that hypothalamic alterations may be more evident in chronic migraine or in more severe migraine phenotypes, whereas direct evidence for consistent macrostructural abnormalities in episodic migraine remains limited. However, this evidence is constrained by the small number of available studies and by the relatively limited sample sizes of episodic migraine cohorts.

In light of converging clinical, preclinical, and functional neuroimaging evidence implicating the hypothalamus in migraine, additional studies are therefore required to determine whether the absence of consistent volumetric findings in episodic migraine reflects a true lack of macrostructural involvement or instead cohort-specific effects related to clinical heterogeneity or methodological differences. Therefore, it is critical to examine whether hypothalamic structural alterations are present in episodic migraine when examined in larger and independent cohorts, whether such alterations represent a reproducible migraine-related feature, and whether they are specific to migraine or instead reflect broader mechanisms shared across pain-related conditions. Longitudinal evidence is also needed to clarify whether hypothalamic volumes remain stable over time or vary in relation to clinical burden.

To address these unresolved questions, the present study applied the same hypothalamic segmentation algorithm employed in recent studies (Xiong et al., 2025; Giardina et al., 2025) to four structural MRI datasets, including two independent cohorts of patients with episodic migraine and two clinical comparison cohorts comprising patients with fibromyalgia or trigeminal neuralgia. Three datasets were publicly available through the OpenNeuro repository, whereas one migraine cohort was acquired as part of our previous work (Ferraro et al., 2026). This design allowed us to test whether episodic migraine is associated with reproducible hypothalamic macrostructural alterations, if they preferentially involve specific hypothalamic subregions, and if they are specific to migraine rather than reflecting a broader structural signature shared across pain-related conditions. In particular, the fibromyalgia dataset was included to assess whether hypothalamic subunit alterations observed in migraine were also present in a chronic widespread pain condition (Clauw, 2014), whereas the trigeminal neuralgia dataset was included to determine whether comparable alterations were present in a chronic craniofacial pain condition involving the trigeminal somatosensory system (Bendtsen et al., 2020). Finally, longitudinal data available for one migraine cohort (Seminowicz et al., 2020) were used to examine the temporal stability of hypothalamic subunit volumes over a follow-up period during which patients underwent two treatment interventions and showed a reduction in headache attack frequency. Together, this approach aimed to clarify the reproducibility, regional specificity, disease specificity, and temporal stability of hypothalamic structural alterations in episodic migraine.

## 2. Methods

### 2.1. Participants

In the present study, we used four independent MRI datasets. These included two cohorts of patients with episodic migraine, namely DS1-MIG, which comprised both baseline data (DS1-MIG-base) and longitudinal follow-up data (DS1-MIG-long) (Seminowicz et al., 2025), and DS2-MIG (Ferraro et al., 2026), as well as two disease-control datasets: one comprising patients with fibromyalgia (DS-FM) (Balducci et al., 2022b) and one comprising patients with trigeminal neuralgia (DS-TN) (Filimonova et al., 2025a). Each dataset included a healthy control (CTRL) group. Except for the DS2-MIG dataset, whose data were acquired in our laboratory, all remaining datasets were obtained from the publicly available OpenNeuro repository (https://openneuro.org). Although this repository contains multimodal MRI data, in the present study, we used only the high-resolution 3D T1-weighted images. Demographic characteristics of the four datasets are reported in Table 1.

**Table 1:** Demographic characteristics of patients and healthy controls across datasets. Age is reported as M ± SD. Sex is reported as number of females/males (F/M). *a*Age range as reported in Seminowicz et al. (2020), which describes a randomized clinical trial including 98 patients with episodic migraine. *b*Age ranges as reported in Filimonova et al. (2025b), which describes 112 patients with trigeminal neuralgia and 48 control individuals. DS1-long is the matched longitudinal subsample of DS1-MIG comprising participants with both baseline and week-20 acquisitions available. All Ns are reported before outlier removal. Abbreviations: MIG = migraine patients; CTRL = healthy controls; FM = fibromyalgia patients; TN = trigeminal neuralgia patients. n.r., not reported in the repository and/or in the paper of reference. * missing information in the repository.

| Dataset | Group | N | Sex (F/M) | Age (years) | Age range | Between-group p-values |
| --- | --- | --- | --- | --- | --- | --- |
| DS1-MIG-base (Migraine) | MIG | 111 | 97 / 14 | n.r. | 18–65<sup>a</sup> | n.r. |
|  | CTRL | 35 | 30 / 4* | n.r. | n.r. |  |
| DS1-MIG-long (Migraine, longitudinal) | MIG | 79 | 73 / 6 | n.r. | 18–65<sup>a</sup> | n.r. |
|  | CTRL | 25 | 23 / 2 | n.r. | n.r. |  |
| DS2-MIG (Migraine) | MIG | 28 | 21 / 7 | 28 ± 8.0 | 18–36 | Sex: $p = .106$; age: $p = .208$ |
| | CTRL | 31 | 17 / 14 | 26 ± 7.2 | 19–47 | |
| DS-FM (Fibromyalgia) | FM | 33 | 33 / 0 | 41.7 ± 6.1 | 30–50 | Age: $p = .887$ |
| | CTRL | 33 | 33 / 0 | 41.5 ± 6.0 | 29–50 | |
| DS-TN (Trigeminal Neuralgia) | TN | 119 | n.r. | n.r. | 30–84<sup>b</sup> | n.r. |
|  | CTRL | 55 | n.r. | n.r. | 38–75<sup>b</sup> |  |

#### Migraine Dataset 1 (DS1-MIG)

The DS1-MIG dataset consisted of MRI data from patients with episodic migraine (MIG) and control participants (CTRL), acquired across three sessions to assess the neural effects of enhanced mindfulness-based stress reduction (MBSR+) and stress management for headache (SMH). The clinical outcomes of these interventions were reported by Seminowicz et al. (2020), showing that both treatments reduced the number of headache days at week 20. However, the reduction was significantly greater in the MBSR+ group (from 7.8 to 4.6 days) compared with the SMH group (from 7.7 to 6.0 days; *p* = .04). Furthermore, 52% of patients receiving MBSR+ achieved a ≥ 50% reduction in headache days, compared with 23% in the SMH group. For this dataset, we performed both cross-sectional and longitudinal analyses. Cross-sectional analyses were based exclusively on the baseline acquisition (DS1-MIG-base), collected before treatment onset. The baseline sample comprised 111 MIG patients (97 females, 14 males) and 35 CTRL participants (30 females, 4 males, 1 participant with missing sex information). For the longitudinal analyses, we included the baseline (DS1-MIG-base) and week-20 acquisitions (DS1-MIG-long), restricting the sample to participants with data available at both time points. This yielded a final longitudinal sample of 79 MIG patients and 25 CTRL participants. For this dataset, participant-level sex information was available in the public repository, whereas participant-level age information was not.

#### Migraine Dataset 2 (DS2-MIG)

The DS2-MIG dataset consisted of MRI data acquired in our laboratory and previously described in Ferraro et al. (2026). Briefly, individuals meeting preliminary eligibility criteria (age 18–65 years, at least one headache attack per month, and headache history longer than one year) were invited for diagnostic classification using the Clinical Decision Support System (CDSS 2.0), an automated interview system with demonstrated high sensitivity and specificity for diagnosing migraine according to the International Classification of Headache Disorders, 3rd edition (ICHD-3) criteria (Han et al., 2023; Headache Classification Committee of the International Headache Society (IHS), 2018). Following CDSS 2.0 classification, 31 patients with MIG were enrolled; 3 were subsequently excluded due to segmentation failure, yielding a final sample of 28 MIG patients (28 ± 8.0 years; 21 females / 7 males). Three patients had migraine with aura; the remaining patients were diagnosed with migraine without aura. None of the included patients reported current or previous use of prophylactic migraine medication. Inclusion criteria for the MIG group required a diagnosis of definite migraine with stable headache characteristics for at least one year prior to study entry. Thirty-one CTRL participants (age, 26 ± 7.2 years; 17 females / 14 males) were enrolled as the comparison group.

Exclusion criteria for all participants included a history of cardiovascular disease, diabetes, hypertension, psychiatric or neurological disorders, and any contraindications to MRI; additional exclusions for migraine patients included comorbidity with other primary or secondary headache disorders. Between-group comparisons showed no significant differences in age or sex distribution. Demographic and clinical characteristics of the DS2-MIG dataset are reported in Table 2. The study was approved by the local ethics committee (School of Life Science and Technology, University of Electronic Science and Technology of China)), and all participants provided written informed consent.

**Table 2:** Demographic and clinical characteristics of the DS2-MIG dataset. The ictal state was defined as having experienced a headache attack within the 72 hours preceding the scanning session. VAS indicates the reported level of pain intensity experienced during headache attacks; NRS indicates the level of pain intensity before the MRI scan; disorder duration indicates the reported years since headache onset; headache frequency indicates the number of attacks per month. Abbreviations: MIG, migraine patients; CTRL, control subjects; VAS, visual analog scale; NRS, numerical rating scale.

|  | MIG (n=28) | CTRL (n= 31) | Statistic | p-value |
| --- | --- | --- | --- | --- |
| Sex (M/F) | 7/21 | 14/17 | $\chi^2(1) = 2.61$ | 0.106 |
| Age (M±SD) | 28±8.0 | 26±7.2 | U=351.0 | 0.208 |
| Disorder duration (years, M±SD) | 9.4±6.99 | - | - | - |
| Headache frequency | 3.1±1.78 | - | - | - |
| Pain level during attacks - VAS: 0-10 (M ± SD) | 6.5±1.55 | - | - | - |
| Pain level before MRI - NRS: 1-9 (M ± SD) | 1.0±0 | - | - | - |
| Patients with an attack in the last 72 hours (yes/no) | 15/13 | - | - | - |
| Patients without aura/with aura | 25/3 | - | - | - |

#### Fibromyalgia Dataset (DS-FM)

The DS-FM dataset consisted of MRI data from 33 FM patients and 33 CTRL participants (Balducci et al., 2022a). FM patients were diagnosed according to the American College of Rheumatology 1990 (Wolfe et al., 1990) and 2016 (Wolfe et al., 2016) criteria. All participants were female, and groups were matched for age (*p* = .887). Participant-level age information was available in the public repository.

#### Trigeminal Neuralgia Dataset (DS-TN)

The DS-TN dataset consisted of MRI data from patients diagnosed with primary trigeminal neuralgia (n = 119) according to ICHD-3 criteria, together with CTRL participants (n = 55) (Filimonova et al., 2025b). Participant-level sex and age information was available in the public repository only for a subset of participants. For the present analyses, only the baseline MRI acquisition was used.

### 2.2. MRI data acquisition

For all datasets, 3D T1-weighted structural images were acquired on 3.0 T MRI scanners, with an isotropic voxel size of 1 × 1 × 1 mm. Detailed scanner, head-coil, and sequence parameters for each dataset are reported in Table SM1. DS1-MIG included two scanner/head-coil configurations, namely a Siemens Tim-Trio scanner equipped with a 32-channel head coil and a Siemens Prisma scanner equipped with a 64-channel head coil. The potential influence of scanner/head-coil configuration was explicitly assessed through sensitivity analyses in both the DS1-MIG cross-sectional and longitudinal analyses (DS1-long).

### 2.3. Structural MRI processing and hypothalamic segmentation

All the 3D T1-weighted images were processed using FreeSurfer (v8.0.0). The standard *recon-all* pipeline was used to derive the total intracranial volume estimate (eTIV, expressed in mm^3^) for each subject. Following the standard processing, hypothalamic segmentations were performed using the dedicated FreeSurfer module (*mri-segment-hypothalamic-subunits*). Importantly, across all datasets, the 3D T1-weighted images were acquired with a spatial resolution of 1×1×1 mm, at the same resolution for which the FreeSurfer segmentation model was optimized (https://surfer.nmr.mgh.harvard.edu/fswiki/HypothalamicSubunit); therefore, no preliminary resampling was required before segmentation.

The *mri-segment-hypothalamic-subunits* algorithm provides probabilistic segmentation of the hypothalamus into 5 bilateral subunits: anterior-inferior, anterior-superior, posterior, tubular-inferior, and tubular-superior (Billot et al., 2020). These labels represent macro-anatomical subdivisions based on histological priors. For each subject, the segmentation pipeline generated a mask for each hypothalamic subunit and extracted the corresponding volume, expressed in mm^3^. These volumetric measures were used in the subsequent statistical analyses, which were performed in R version 4.5.3.

### 2.4. Hypothalamic subunit volume adjustment for estimated intracranial volume (eTIV)

To minimize the influence of global head-size differences on regional hypothalamic volumes, all volumetric measures were adjusted for estimated total intracranial volume (eTIV) using the residuals method (Sanchis-Segura et al.; Mathalon et al.; Voevodskaya et al., 2014).

This approach removes variance in regional volume linearly attributable to eTIV and is considered an appropriate strategy for intracranial-volume correction in volumetric MRI analyses, reducing apparent sex-related gray-matter differences after adjustment (Sanchis-Segura et al.; Voevodskaya et al., 2014).

For each dataset and subunit, residualization was performed by estimating the eTIV correction model in CTRL participants only; the resulting model was then applied to all participants (Voevodskaya et al., 2014). Specifically adjusted volumes were computed as follows:

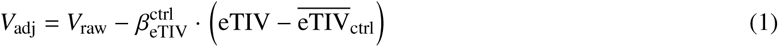

where 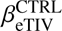 is the ordinary least-squares regression slope of the subunit volume on eTIV estimated within the control group, and 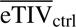 is the mean eTIV of the control group. The same control-derived 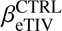 was then applied to all participants (both patients and CTRL) to compute *V*_adj_, ensuring that the volume–eTIV scaling relationship was not contaminated by disease-related variance.

Bilateral subunit volumes were subsequently obtained as the sum of the left and right subunits adjusted values.

This procedure has two important properties: (i) the resulting adjusted values remain interpretable on the original volumetric scale (mm^3^), rather than as standardized residuals, facilitating comparison with raw values and other studies; and (ii) deriving the adjustment slope from the CTRL group reduces the risk that disease-related volumetric variance is partially absorbed into the eTIV correction term, as may occur when the regression is fitted on the pooled patient-control sample (Voevodskaya et al., 2014).

### 2.5. Outlier detection

To identify and address potentially aberrant volumetric measurements while preserving statistical power, we implemented a hybrid outlier strategy separately within each dataset. Outlier detection was performed on eTIV-adjusted volumes using Tukey’s 1.5 × IQR rule and was applied independently to each eTIV-adjusted hypothalamic subunit volume included in the subsequent analyses. Importantly, no cross-variable propagation was performed: an outlier flagged on the bilateral measure of a subunit did not automatically result in flagging of the corresponding left or right hemispheric measures, and vice versa.

For each subject, we tabulated the number of distinct anatomical subunits flagged as outliers. Here, “root subunit” refers to the anatomical structure irrespective of laterality or aggregation level; for example, a subject flagged on the bilateral, left, or right anterior-superior volume was considered to have one affected root subunit.

We then applied a two-tier decision rule. First, multi-unit outliers, defined as subjects flagged on two or more distinct root subunits, were considered likely to reflect segmentation failure or systematic measurement error and were therefore excluded entirely from the corresponding dataset. Second, single-unit outliers, defined as subjects flagged on only one root subunit irrespective of whether the flagged value involved the bilateral, left, and/or right measure, were retained. For these subjects, only the specific flagged value(s) were set to NA, with no propagation to non-flagged measures of the same subunit or to other hypothalamic subunits.

This hybrid strategy is more conservative than complete subject exclusion (which would discard otherwise valid information) and more robust than retaining all values (which could allow a single mis-segmented voxel to drive group-level effects).

### 2.6. Sensitivity analyses of residual sex- and age-related effects after eTIV adjustment in CTRL groups

Because participant-level demographic information was incomplete across datasets, formal covariate-based analyses, including age and sex, could not be applied uniformly to all samples. Therefore, before conducting the main group analyses, we performed sensitivity analyses restricted to CTRL participants from datasets with available demographic information to evaluate whether eTIV residualization reduced demographic-related variance in hypothalamic subunit volumes.

Sex effects were assessed in CTRL participants from DS1-MIG-base and DS2-MIG, as these were the only CTRL samples including both male and female participants with available participant-level sex information. Raw and eTIV-adjusted bilateral hypothalamic subunit volumes were compared between sexes using Welch’s t-tests.

Age effects were assessed in CTRL participants from DS2-MIG and DS-FM, for which participant-level age information was available. Associations between age and raw or eTIV-adjusted bilateral hypothalamic subunit volumes were evaluated using Spearman’s rho.

### 2.7. Cross-sectional group-comparison analyses

In each dataset (DS1-MIG-base, DS2-MIG, DS-FM, and DS-TN), the main cross-sectional analyses followed a hierarchical decomposition framework.

First, group differences (patients vs. CTRL) were tested at the bilateral anatomical level using an omnibus MANOVA including the 5 bilateral eTIV-adjusted hypothalamic subunit volumes as dependent variables. This multivariate test was used to assess whether each dataset showed an overall group effect across hypothalamic subunits. The main analysis was completed with several sensitivity analyses.

Second, bilateral effects were decomposed using univariate group comparisons for each eTIV-adjusted bilateral subunit volume.

Third, to further characterize the spatial pattern of any group differences and assess potential hemispheric specificity, the same univariate analyses were then performed separately for the corresponding left- and right-hemisphere eTIV-adjusted subunit volumes.

Multivariate test statistics were extracted using summary.manova with statistical significance defined as *p* < .05.

#### 2.7.1. Bilateral hypothalamic subunit volumes: multivariate analyses (MANOVAs)

To test whether patients differed from CTRL participants in the overall bilateral hypothalamic subunit volume profile, we conducted a one-way MANOVA separately within each dataset. The 5 bilateral eTIV-adjusted hypothalamic subunit volumes (anterior-inferior, anterior-superior, posterior, tubular-inferior, and tubular-superior) were included as dependent variables, with group as the between-subjects factor. Pillai’s trace was used as the primary multivariate statistic because of its robustness to mild violations of multivariate normality and homogeneity of covariance matrices (Olson, 1976).

MANOVA was conducted using listwise deletion: participants with missing values in any of the 5 bilateral dependent variables were excluded from the multivariate test. Thus, in the primary cleaned dataset, subjects with single-unit outliers affecting one or more bilateral measures were excluded from the bilateral MANOVA. Distributional assumptions were inspected using per-group Q-Q plots for each subunit. Minor deviations from normality were observed in some anterior subunits, but no transformation was applied given the relative robustness of Pillai’s trace.

##### Sensitivity analysis for outlier handling

To assess robustness to outlier handling, the MANOVA was repeated also using the full uncleaned sample without outlier removal. Omnibus results were compared across cleaning levels to evaluate the sensitivity of the multivariate group effect to outlier-handling choices.

##### Sex- and age-adjusted sensitivity analysis

As an additional sensitivity analysis, and only in datasets where participant-level sex (DS1-MIG-base and DS2-MIG) and age information were available (DS2-MIG), we also conducted a MANCOVA including sex as categorical factor and age as a continuous covariate. This analysis was used to assess whether the multivariate group effect was robust to adjustment for sex, and the contribution of sex was reported as a separate term in the model.

##### Coil-homogeneous sensitivity analysis in DS1-MIG

To assess whether the DS1-MIG multivariate findings were influenced by the use of different head coils, we performed a coil-homogeneous sensitivity analysis restricted to subjects acquired with Coil 1 only. The same 5 bilateral eTIV-adjusted hypothalamic subunit volumes were entered as dependent variables in a one-way MANOVA with group (MIG vs. CTRL) as the between-subjects factor. The primary Coil sensitivity analysis followed the same hybrid outlier-handling strategy used in the main analysis.

##### Peri-ictal sensitivity analysis in DS2-MIG

To assess whether the DS2-MIG multivariate group effect was driven by MIG patients scanned in temporal proximity to a migraine attack, we performed an additional sensitivity analysis. Migraineurs were classified according to the number of days since their last migraine attack at the time of MRI acquisition, while CTRL participants were retained in all analyses. The sensitivity model excluded migraineurs scanned within 3 days of their last attack before MRI acquisition, according to the criterion used by Giardina et al. (2025). This analysis was restricted to the omnibus multivariate effect and was used to evaluate whether the main bilateral hypothalamic group effect was robust to the exclusion of peri-ictal patients.

#### 2.7.2. Bilateral hypothalamic subunit volumes: univariate follow-up analyses

Following the bilateral hypothalamic subunit volumes omnibus MANOVA, we performed univariate decomposition on each of the 5 bilateral eTIV-adjusted subunits to identify which structures contributed to the multivariate group effect. For each subunit, Welch’s t-test (patients vs. CTRL) was used as the primary test, complemented by a Mann–Whitney U test as a non-parametric robustness check. Effect sizes were quantified using Hedges’ *g* with 95% confidence intervals (Hedges, 1981).

Univariate tests were performed on pairwise-complete observations, excluding only the specific missing value for each subunit. Benjamini–Hochberg FDR correction was applied across the 5 bilateral subunits; significance was set at adjusted *p* < .05.

#### 2.7.3. Left- and right-hemisphere subunit volumes: univariate follow-up analyses

The same univariate framework was applied separately to the left- and right-hemisphere eTIV-adjusted subunit volumes to assess whether group differences showed hemispheric specificity. Specifically, we tested the 10 side-specific eTIV-adjusted hypothalamic subunit volumes. For each side-specific subunit, Welch’s two-sample t-test was used as the primary group comparison, given its robustness to unequal sample sizes and heteroscedasticity. Effect sizes were quantified using Hedges’ *g* with 95% confidence intervals. Analyses were performed using pairwise-complete observations, excluding only missing values for the specific subunit being tested.

To control for multiple comparisons, p-values were adjusted using the Benjamini–Hochberg FDR procedure at two levels. First, within-side FDR correction was applied separately within each hemisphere across the 5 subunit tests. Second, as a more conservative sensitivity correction, FDR adjustment was also applied across all 10 side-specific subunit tests within each cohort. Findings surviving within-side FDR correction were interpreted as hemisphere-specific effects, whereas findings additionally surviving the all-10 correction were considered the most robust side-specific effects. The side-specific analyses were used as localized anatomical follow-ups to characterize the hemispheric distribution of group differences observed at the bilateral level.

### 2.8. Longitudinal analyses on dataset DS1-MIG

To examine whether the hypothalamic volumetric abnormality identified in the baseline univariate analyses persisted after treatment-related reductions in headache frequency, we conducted longitudinal follow-up analyses in the subset of DS1-MIG participants with both baseline (DS1-MIG-base) and week-20 follow-up (DS1-MIG-long) MRI acquisitions. This yielded a matched analytic sample of 99 participants (24 CTRL and 75 migraineurs).

The primary longitudinal outcome was defined as the hypothalamic subunit volume that emerged as significant in the baseline univariate analyses. For this outcome, we fitted a linear mixed-effects model testing the group-by-time interaction:

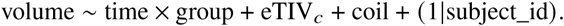

In this model, *time* indexed baseline and week-20 follow-up, *group* indexed MIG versus CTRL, baseline eTIV centered on the CTRL mean was included as a between-subjects covariate, and acquisition coil (Coil1/Coil2) was included as a time-varying covariate because 12 of 99 matched participants changed coil between acquisitions. The main effect of interest was the time × group interaction, testing whether longitudinal volume changes differed between MIG patients and CTRL participants.

For these longitudinal models, raw, non-residualized hypothalamic volumes were used as dependent variables. This choice reflected a methodological distinction from the cross-sectional analyses: because intracranial volume is stable over time, applying separate control-derived residualization slopes at each time point could introduce artificial between-timepoint shifts unrelated to biological change. Instead, baseline eTIV was included directly as a covariate. To verify robustness to coil changes, a sensitivity analysis excluding the 12 participants who changed coil between baseline and follow-up was conducted, yielding a reduced matched sample of 87 participants. As a secondary analysis, the same group-by-time model was applied to all 10 side-specific hypothalamic subunit volumes, with p-values adjusted using the Benjamini–Hochberg FDR procedure. As an exploratory analysis within the MIG group, we additionally tested whether the two treatment arms (MBSR+ vs. SMH) differed in their longitudinal trajectories on the primary outcome.

Models were estimated by restricted maximum likelihood using lme4/lmerTest, with Satterthwaite degrees of freedom. Effect sizes were quantified as Hedges’ *g*_av_ for paired observations.

## 3. Results

### 3.1. Sensitivity analyses of residual sex- and age-related effects after eTIV adjustment in CTRL groups

For the results of this section, see Table SM2. Before eTIV correction, raw volumes showed nominal sex-related differences in several hypothalamic subunits, particularly in DS2-MIG, although none reached conventional statistical significance (*p*< .05). Following eTIV adjustment, no subunit showed a significant sex difference in either dataset (all adjusted p-values *p* = .145 in DS1-MIG-base and > .242 in DS2-MIG). These findings suggest that eTIV residualization substantially reduced sex-related volumetric scaling effects in the CTRL samples.

Age-related associations were limited. In the raw volumes, no systematic correlations between age and hypothalamic subunit volumes were observed in the CTRL samples with available age information. The only nominal association emerged in DS2-MIG, where tubular-inferior volume showed a weak negative correlation with age (*ρ* = −.373, *p* = .039). This association was attenuated after eTIV adjustment and no longer reached conventional significance (*ρ* = −.334, *p* = .067). In the DS-FM CTRL sample, no hypothalamic subunit showed a significant association with age either before or after eTIV adjustment (all *p*> .13).

### 3.2. Bilateral hypothalamic subunit volumes: multivariate analyses (MANOVAs)

For the results of this section, see Tables 3 and 4. In DS1-MIG-base, the omnibus MANOVA on the 5 bilateral eTIV-adjusted hypothalamic subunits revealed a significant multivariate effect of group (MIG vs. CTRL) in the primary analysis [F(5, 124) = 3.43, *p* = .006]. Inclusion of biological sex as a covariate preserved the group effect [F(5, 123) = 3.40, *p* = .007], while sex itself did not significantly contribute to the multivariate model.

**Table 3:** eTIV-adjusted hypothalamic subunit volumes (mm^3^) by group and dataset. Values are expressed as M ± SD. Whole hypothalamus is the sum of the five bilateral subunits and is reported as an exploratory measure (outside the FDR family). eTIV-adjusted subunit measures computed using the residualization method (controls-only *β*) in each dataset. Sample sizes vary by subunit due to hybrid outlier handling (single-unit outlier values set to NA). Volumes are not directly comparable across datasets due to scanner-dependent absolute scaling. Abbreviations: CTRL = healthy controls; MIG = migraine patients; FM = fibromyalgia patients; eTIV= estimated total intracranial volume; TN = trigeminal neuralgia patients; ANT-INF = anterior inferior; ANT-SUP = anterior superior; POST = posterior; TUB-INF = tubular inferior; TUB-SUP = tubular superior.

| Subunit | DS1-MIG-base |  | DS2-MIG |  | DS-FM |  | DS-TN |  |
| --- | --- | --- | --- | --- | --- | --- | --- | --- |
| | CTRL ($n=35$) | MIG ($n=103-108$) | CTRL ($n=29-31$) | MIG ($n=24-27$) | CTRL ($n=32-33$) | FM ($n=30-32$) | CTRL ($n=49-52$) | TN ($n=106-108$) |
| ANT-INF | $38.4 \pm 4.0$ | $37.5 \pm 4.5$ | $19.0 \pm 5.3$ | $23.9 \pm 6.2$ | $33.9 \pm 6.1$ | $33.4 \pm 5.2$ | $26.0 \pm 7.1$ | $25.0 \pm 6.7$ |
| ANT-SUP | $49.9 \pm 4.6$ | $51.7 \pm 6.1$ | $37.0 \pm 4.2$ | $41.1 \pm 5.1$ | $48.2 \pm 7.3$ | $46.5 \pm 6.4$ | $39.0 \pm 5.4$ | $37.9 \pm 6.4$ |
| POST | $229.8 \pm 19.4$ | $238.7 \pm 18.7$ | $201.1 \pm 19.3$ | $203.7 \pm 22.4$ | $193.9 \pm 22.1$ | $191.7 \pm 16.1$ | $205.7 \pm 20.1$ | $203.9 \pm 22.1$ |
| TUB-INF | $266.8 \pm 21.1$ | $264.1 \pm 20.7$ | $229.6 \pm 33.7$ | $244.1 \pm 23.7$ | $240.0 \pm 19.1$ | $241.7 \pm 24.3$ | $240.6 \pm 16.7$ | $244.7 \pm 29.3$ |
| TUB-SUP | $234.5 \pm 17.6$ | $237.0 \pm 21.6$ | $222.4 \pm 14.4$ | $223.6 \pm 15.0$ | $211.1 \pm 16.6$ | $217.4 \pm 12.6$ | $214.4 \pm 18.5$ | $212.5 \pm 16.5$ |
| Whole | $820.5 \pm 49.3$ | $829.7 \pm 50.6$ | $708.4 \pm 58.4$ | $736.0 \pm 53.7$ | $726.6 \pm 53.2$ | $727.9 \pm 46.7$ | $726.2 \pm 50.3$ | $722.2 \pm 59.1$ |

**Table 4:** Multivariate analysis of bilateral hypothalamic subunit volumes across the different datasets. For each dataset, the five bilateral subunit volumes were tested jointly. MANOVA results for full uncleaned and primary outlier-handled models are shown to evaluate sensitivity to outlier handling. MANCOVA models include biological sex as a categorical covariate where applicable; “group | sex” denotes the group effect adjusted for sex, and “sex | group” denotes the sex effect adjusted for group. “group | age” and “age | group” denote the corresponding effects when age is additionally included as a continuous covariate. MANCOVA was not performed for DS-FM because all participants were female nor for DS-TN. After all, participant-level sex information was not available for the full analytic sample. Abbreviations: CTRL = healthy controls; PAT = patients; eTIV= estimated total intracranial volume; ANT-INF = anterior inferior; ANT-SUP = anterior superior; POST = posterior; TUB-INF = tubular inferior; TUB-SUP = tubular superior.

| Cohort | Model | n (CTRL/PAT) | Pillai | F(df) | p |
| --- | --- | --- | --- | --- | --- |
| DS1-MIG-base | MANOVA, full uncleaned | 145 (34/111) | 0.093 | 2.86 (5, 139) | .017 |
|  | MANOVA, primary cleaned | 130 (31/99) | 0.121 | 3.43 (5, 124) | .006 |
|  | MANCOVA, group \| sex | 130 (31/99) | 0.121 | 3.40 (5, 123) | .007 |
|  | MANCOVA, sex \| group | 130 (31/99) | 0.030 | 0.75 (5, 123) | .587 |
| DS2-MIG | MANOVA, full uncleaned | 59 (31/28) | 0.161 | 2.03 (5, 53) | .089 |
|  | MANOVA, primary cleaned | 52 (28/24) | 0.283 | 3.63 (5, 46) | .008 |
|  | MANCOVA, group \| sex | 52 (28/24) | 0.298 | 3.82 (5, 45) | .006 |
|  | MANCOVA, sex \| group | 52 (28/24) | 0.035 | 0.33 (5, 45) | .894 |
|  | MANCOVA, group \| sex, age | 52 (28/24) | 0.336 | 4.44 (5, 44) | .002 |
|  | MANCOVA, age \| sex, group | 52 (28/24) | 0.117 | 1.16 (5, 44) | .343 |
| DS-FM | MANOVA, full uncleaned | 66 (33/33) | 0.019 | 0.23 (5, 60) | .950 |
|  | MANOVA, primary cleaned | 61 (31/30) | 0.084 | 1.01 (5, 55) | .418 |
|  | MANCOVA, group \| age | 61 (31/30) | 0.088 | 1.04 (5, 54) | .404 |
|  | MANCOVA, age \| group | 61 (31/30) | 0.099 | 1.19 (5, 54) | .325 |
| DS-TN | MANOVA, full uncleaned | 159 (49/110) | 0.017 | 0.52 (5, 153) | .760 |
|  | MANOVA, primary cleaned | 143 (42/101) | 0.043 | 1.22 (5, 137) | .303 |

Subject-specific age was not available for the whole dataset and could therefore not be included as a covariate.

In DS2-MIG the omnibus MANOVA also showed a significant multivariate effect of group in the primary analysis [F(5, 46) = 3.63, *p* = .008]. Inclusion of biological sex as a covariate preserved and slightly strengthened the group effect [F(5, 45) = 3.82, *p* = .006], while sex itself did not significantly contribute to the model. Further inclusion of age as an additional covariate yielded a comparable pattern: the group effect was further strengthened [F(5, 44) = 4.44, *p* = .002], while neither sex nor age reached significance as independent predictors (both *p* > .34), confirming that these variables did not confound the observed group differences.

In the DS-FM dataset, the omnibus MANOVA revealed no significant multivariate group effect (FM vs. CTRL) in the primary analysis. Given the absence of sex variation (all participants female), no MANCOVA with sex as a covariate was performed.

In the DS-TN dataset, the omnibus MANOVA revealed no significant multivariate effect of group (TN vs. CTRL) in the primary analysis. Since all participants were female, sex was not included as a covariate. Age was well-matched between groups and its inclusion as a covariate did not alter the result.

Overall, the omnibus multivariate analyses revealed a significant group effect on bilateral hypothalamic subunit volumes in both migraine cohorts (DS1-MIG and DS2-MIG), but not in the fibromyalgia (DS-FM) or trigeminal neuralgia (DS-TN) disease-control cohorts. The DS1-MIG effect was robust across outlier-handling strategies and after adjustment for biological sex, whereas the DS2-MIG effect was significant in the primary cleaned analysis and remained significant after sex adjustment, although it was only directionally present in the full uncleaned sample.

*Coil-homogeneous sensitivity analysis in DS1-MIG-base.* In the coil-homogeneous sensitivity analysis in the dataset DS1-MIG-base, using the same hybrid outlier-handling and listwise-deletion procedure as in the main analysis, the multivariate group effect remained significant [F(5, 112) = 3.21, *p* = .010] (see Table SM3). Thus, the DS1-MIG-base omnibus effect was not abolished when the analysis was restricted to a coil-homogeneous subset.

*Peri-ictal sensitivity analysis in DS2-MIG.* In DS2-MIG, after excluding migraineurs whose last attack occurred less than 3 days before MRI acquisition, the multivariate group effect remained significant and numerically increased in magnitude [28 CTRL, 15 MIG; F(5, 37) = 4.09, *p* = .005] (see Table SM4). The effect was similarly preserved after adjustment for sex [F(5, 36) = 4.25, *p* = .004], with no significant contribution of sex.

### 3.3. Bilateral hypothalamic subunit volumes: univariate follow-up analysis

For the results of this section, see Table 5. In DS1-MIG-base, univariate decomposition did not reveal any FDR-significant group difference across the 5 bilateral hypothalamic subunits.

**Table 5:** Univariate decomposition of the bilateral MANOVA. Welch’s two-sample *t*-test on each of the five eTIV-adjusted hypothalamic subunits (patients vs. control subjects) with Hedges’ *g* effect sizes and 95% confidence intervals. Effect sizes are calculated using the small-sample correction factor *J* = 1 − 3/[4(*n*_1_ + *n*_2_) − 9]. *p*-values are adjusted for multiple comparisons across the five sub-units within each cohort using the Benjamini-Hochberg FDR procedure. Pairwise-complete observations were used (sample sizes vary by sub-unit due to outlier-driven NA-setting). Positive *g*indicates larger volume in patients than control subjects. Abbreviations: CTRL = healthy controls; PAT = patients

| Cohort | Subunit | n (CTRL/PAT) | Hedges’ $g$ | 95% CI | $p_{\text{Welch}}$ | $p_{\text{FDR}}$ |
| --- | --- | --- | --- | --- | --- | --- |
| DS1-MIG-base | Anterior-inferior | 33/103 | −0.21 | [−0.60, +0.18] | .268 | .446 |
|  | Anterior-superior | 33/107 | +0.31 | [−0.09, +0.69] | .078 | .195 |
|  | Posterior | 35/106 | +0.47 | [+0.09, +0.86] | .020 | .101 |
|  | Tubular-inferior | 35/107 | −0.13 | [−0.51, +0.25] | .517 | .517 |
|  | Tubular-superior | 35/108 | +0.12 | [−0.26, +0.50] | .501 | .517 |
| DS2-MIG | Anterior-inferior | 31/27 | +0.84 | [+0.30, +1.37] | .003 | .006 |
|  | Anterior-superior | 29/26 | +0.86 | [+0.31, +1.41] | .002 | .006 |
|  | Posterior | 31/26 | +0.12 | [−0.39, +0.64] | .642 | .757 |
|  | Tubular-inferior | 31/27 | +0.49 | [−0.03, +1.00] | .061 | .101 |
|  | Tubular-superior | 30/26 | +0.08 | [−0.44, +0.60] | .757 | .757 |
| DS-FM | Anterior-inferior | 33/32 | −0.09 | [−0.57, +0.39] | .706 | .751 |
|  | Anterior-superior | 33/32 | −0.25 | [−0.73, +0.24] | .319 | .751 |
|  | Posterior | 32/32 | −0.11 | [−0.60, +0.37] | .649 | .751 |
|  | Tubular-inferior | 33/32 | +0.08 | [−0.40, +0.56] | .751 | .751 |
|  | Tubular-superior | 32/30 | +0.42 | [−0.08, +0.91] | .101 | .504 |
| DS-TN | Anterior-inferior | 45/105 | −0.34 | [−0.69, +0.01] | .040 | .198 |
|  | Anterior-superior | 52/110 | −0.20 | [−0.53, +0.13] | .203 | .440 |
|  | Posterior | 52/107 | −0.13 | [−0.46, +0.20] | .440 | .440 |
|  | Tubular-inferior | 49/109 | +0.13 | [−0.21, +0.46] | .360 | .440 |
|  | Tubular-superior | 52/111 | −0.15 | [−0.47, +0.18] | .399 | .440 |

In DS2-MIG, univariate decomposition identified significantly larger anterior-inferior and anterior-superior volumes in MIG patients than in CTRL participants, with both effects surviving FDR correction (both *p*_FDR_ = .006).

In the DS-FM dataset, univariate decomposition revealed no FDR-significant group differences across the 5 bilateral hypothalamic subunits, nor any uncorrected effect reaching *p* < .05. Effect sizes were small to moderate, and all 95% confidence intervals crossed zero.

In DS-TN, univariate decomposition revealed no significant group differences across the 5 bilateral eTIV-adjusted hypothalamic subunits, either before or after FDR correction. Effect sizes were small, and all 95% confidence intervals crossed zero.

### 3.4. Left- and right-hemisphere subunit volumes: univariate follow-up analysis

For the results of this section, see Table 6. To further characterize the hemisphere-specific anatomical expression of the bilateral multivariate group effect, we examined the 10 side-specific eTIV-adjusted hypothalamic subunit volumes across the different datasets.

**Table 6:**
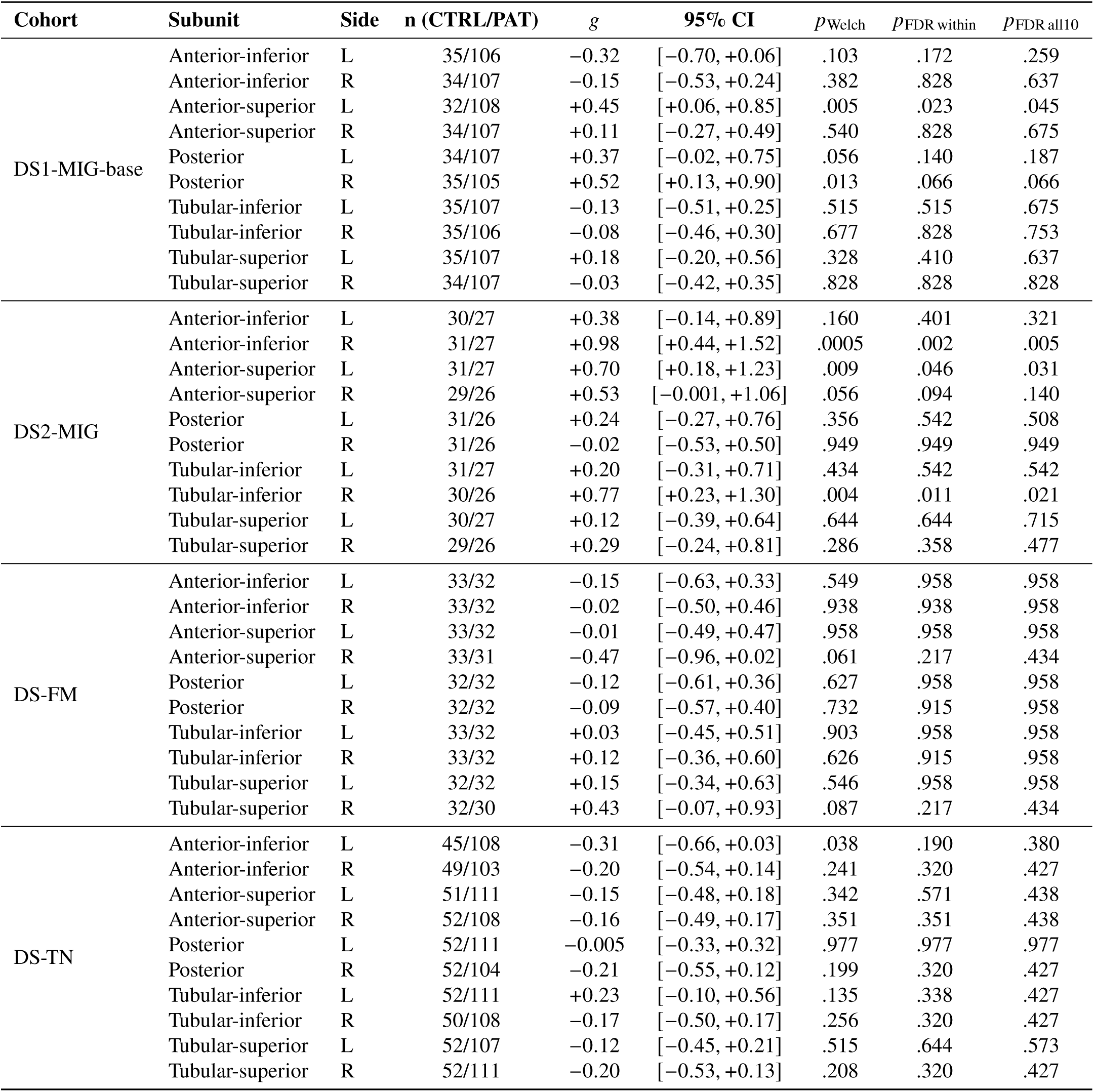
Per-side univariate decomposition of the lateralized MANOVAs. Welch’s *t*-test on each ICV-adjusted hypothalamic sub-unit by hemisphere, with Hedges’ *g*effect sizes (patients vs. Control; positive *g* indicates larger volume in patients) and 95% confidence intervals. *p*_FDR within_ is corrected within hemisphere (5 tests per family); *p*_FDR all10_ is corrected across all 10 lateralized variables (sensitivity).

| Cohort | Subunit | Side | n (CTRL/PAT) | $g$ | 95% CI | $p_{\text{Welch}}$ | $p_{\text{FDR within}}$ | $p_{\text{FDR all10}}$ |
| --- | --- | --- | --- | --- | --- | --- | --- | --- |
| DS1-MIG-base | Anterior-inferior | L | 35/106 | -0.32 | [-0.70, +0.06] | .103 | .172 | .259 |
|  | Anterior-inferior | R | 34/107 | -0.15 | [-0.53, +0.24] | .382 | .828 | .637 |
|  | Anterior-superior | L | 32/108 | +0.45 | [+0.06, +0.85] | .005 | .023 | .045 |
|  | Anterior-superior | R | 34/107 | +0.11 | [-0.27, +0.49] | .540 | .828 | .675 |
|  | Posterior | L | 34/107 | +0.37 | [-0.02, +0.75] | .056 | .140 | .187 |
|  | Posterior | R | 35/105 | +0.52 | [+0.13, +0.90] | .013 | .066 | .066 |
|  | Tubular-inferior | L | 35/107 | -0.13 | [-0.51, +0.25] | .515 | .515 | .675 |
|  | Tubular-inferior | R | 35/106 | -0.08 | [-0.46, +0.30] | .677 | .828 | .753 |
|  | Tubular-superior | L | 35/107 | +0.18 | [-0.20, +0.56] | .328 | .410 | .637 |
|  | Tubular-superior | R | 34/107 | -0.03 | [-0.42, +0.35] | .828 | .828 | .828 |
| DS2-MIG | Anterior-inferior | L | 30/27 | +0.38 | [-0.14, +0.89] | .160 | .401 | .321 |
|  | Anterior-inferior | R | 31/27 | +0.98 | [+0.44, +1.52] | .0005 | .002 | .005 |
|  | Anterior-superior | L | 31/27 | +0.70 | [+0.18, +1.23] | .009 | .046 | .031 |
|  | Anterior-superior | R | 29/26 | +0.53 | [-0.001, +1.06] | .056 | .094 | .140 |
|  | Posterior | L | 31/26 | +0.24 | [-0.27, +0.76] | .356 | .542 | .508 |
|  | Posterior | R | 31/26 | -0.02 | [-0.53, +0.50] | .949 | .949 | .949 |
|  | Tubular-inferior | L | 31/27 | +0.20 | [-0.31, +0.71] | .434 | .542 | .542 |
|  | Tubular-inferior | R | 30/26 | +0.77 | [+0.23, +1.30] | .004 | .011 | .021 |
|  | Tubular-superior | L | 30/27 | +0.12 | [-0.39, +0.64] | .644 | .644 | .715 |
|  | Tubular-superior | R | 29/26 | +0.29 | [-0.24, +0.81] | .286 | .358 | .477 |
| DS-FM | Anterior-inferior | L | 33/32 | -0.15 | [-0.63, +0.33] | .549 | .958 | .958 |
|  | Anterior-inferior | R | 33/32 | -0.02 | [-0.50, +0.46] | .938 | .938 | .958 |
|  | Anterior-superior | L | 33/32 | -0.01 | [-0.49, +0.47] | .958 | .958 | .958 |
|  | Anterior-superior | R | 33/31 | -0.47 | [-0.96, +0.02] | .061 | .217 | .434 |
|  | Posterior | L | 32/32 | -0.12 | [-0.61, +0.36] | .627 | .958 | .958 |
|  | Posterior | R | 32/32 | -0.09 | [-0.57, +0.40] | .732 | .915 | .958 |
|  | Tubular-inferior | L | 33/32 | +0.03 | [-0.45, +0.51] | .903 | .958 | .958 |
|  | Tubular-inferior | R | 33/32 | +0.12 | [-0.36, +0.60] | .626 | .915 | .958 |
|  | Tubular-superior | L | 32/32 | +0.15 | [-0.34, +0.63] | .546 | .958 | .958 |
|  | Tubular-superior | R | 32/30 | +0.43 | [-0.07, +0.93] | .087 | .217 | .434 |
| DS-TN | Anterior-inferior | L | 45/108 | -0.31 | [-0.66, +0.03] | .038 | .190 | .380 |
|  | Anterior-inferior | R | 49/103 | -0.20 | [-0.54, +0.14] | .241 | .320 | .427 |
|  | Anterior-superior | L | 51/111 | -0.15 | [-0.48, +0.18] | .342 | .571 | .438 |
|  | Anterior-superior | R | 52/108 | -0.16 | [-0.49, +0.17] | .351 | .351 | .438 |
|  | Posterior | L | 52/111 | -0.005 | [-0.33, +0.32] | .977 | .977 | .977 |
|  | Posterior | R | 52/104 | -0.21 | [-0.55, +0.12] | .199 | .320 | .427 |
|  | Tubular-inferior | L | 52/111 | +0.23 | [-0.10, +0.56] | .135 | .338 | .427 |
|  | Tubular-inferior | R | 50/108 | -0.17 | [-0.50, +0.17] | .256 | .320 | .427 |
|  | Tubular-superior | L | 52/107 | -0.12 | [-0.45, +0.21] | .515 | .644 | .573 |
|  | Tubular-superior | R | 52/111 | -0.20 | [-0.53, +0.13] | .208 | .320 | .427 |

In DS1-MIG-base, the left anterior-superior subunit was significantly larger in MIG patients than in CTRL participants and survived both within-side and all-10 FDR correction (within-side *p*_FDR_ = .023; all-10 *p*_FDR_ = .045). No remaining left- or right-hemisphere subunit survived FDR correction.

In DS2-MIG, three side-specific subunits survived both within-side and all-10 FDR correction, all showing larger volumes in MIG patients than in CTRL participants. These included the right anterior-inferior subunit (within-side *p*_FDR_ = .002; all-10 *p*_FDR_ = .005), the right tubular-inferior subunit (within-side *p*_FDR_ = .011; all-10 *p*_FDR_ = .021), and the left anterior-superior subunit (within-side *p*_FDR_ = .046; all-10 *p*_FDR_ = .031). No other side-specific subunit survived FDR correction.

In DS-FM, no side-specific hypothalamic subunit showed a significant group difference after either within-hemisphere or all-10 FDR correction. Results were non-significant for all subunits in both hemispheres, and all confidence intervals crossed zero.

Similarly, in DS-TN, no side-specific hypothalamic subunit showed a significant group difference after either within-hemisphere or all-10 FDR correction. All confidence intervals crossed zero.

### 3.5. Longitudinal analyses on the DS1-MIG dataset

For the results of this section, see Table 7. The primary longitudinal analysis revealed no significant group-by-time interaction for the left anterior-superior subunit (*p* = .754), the pre-specified primary outcome.

**Table 7:** Longitudinal analysis of lateralized hypothalamic sub-unit volumes in DS1-MIG (DS1-MIG=base and DS1-MIG-long). Linear mixed model: *volume* ∼ *time* × *group* + TIV*c* + *coil* + (1|*subject*), estimated on *n* = 99 matched pairs (24 controls, 75 migraineurs; follow-up interval: acq1 → acq3). Hedges’ *g*_av_ quantifies paired within-group change (follow-up − baseline); positive values indicate volume increase over time. FDR correction (Benjamini–Hochberg) applied across all 10 lateralized sub-units for the group × time interaction term. The left anterior-superior subunit was the pre-specified primary region of interest; therefore, no FDR correction was applied to this test. Sensitivity analysis excluding the 12 participants who changed the acquisition coil between timepoints (*n* = 87: 23 controls, 64 migraineurs) yielded *p*_interaction_ = .873, *g*^Mig^ = +0.11 [−0.13, +0.36], confirming robustness to the coil change.

| Sub-unit | Side | $n$ (CTRL/MIG) | $g_{av}$ Migraine | | $g_{av}$ Control | | $p_{interaction}$ | $p_{FDR}$ |
| --- | --- | --- | --- | --- | --- | --- | --- | --- |
| | | | $g_{av}$ | 95% CI | $g_{av}$ | 95% CI | | |
| *Primary outcome (pre-specified, no FDR)* |  |  |  |  |  |  |  |  |
| Anterior-superior | L | 24/75 | +0.09 | [−0.13, +0.32] | +0.18 | [−0.23, +0.58] | .754 | — |
| *Secondary outcomes (FDR corrected across 10 lateralized sub-units)* |  |  |  |  |  |  |  |  |
| Anterior-inferior | L | 24/75 | +0.08 | [−0.15, +0.30] | +0.13 | [−0.28, +0.53] | .977 | .977 |
| Anterior-inferior | R | 24/75 | +0.16 | [−0.06, +0.39] | −0.09 | [−0.49, +0.31] | .195 | .651 |
| Anterior-superior | R | 24/75 | +0.07 | [−0.16, +0.29] | −0.04 | [−0.44, +0.36] | .570 | .815 |
| Posterior | L | 24/75 | −0.14 | [−0.37, +0.09] | +0.21 | [−0.20, +0.61] | .007 | .068 |
| Posterior | R | 24/75 | −0.10 | [−0.33, +0.12] | +0.08 | [−0.32, +0.48] | .170 | .651 |
| Tubular-inferior | L | 24/75 | +0.07 | [−0.15, +0.30] | +0.07 | [−0.33, +0.47] | .904 | .977 |
| Tubular-inferior | R | 24/75 | +0.05 | [−0.17, +0.28] | +0.19 | [−0.22, +0.59] | .421 | .751 |
| Tubular-superior | L | 24/75 | −0.12 | [−0.35, +0.10] | −0.32 | [−0.73, +0.09] | .451 | .751 |
| Tubular-superior | R | 24/75 | −0.12 | [−0.35, +0.11] | +0.04 | [−0.36, +0.45] | .338 | .751 |
| *Exploratory: treatment-arm within-group changes on ANT-SUP L (migraineurs only)* |  |  |  |  |  |  |  |  |
| MBSR+ ($n = 37$) | | | $\Delta = +0.07 \text{ mm}^3$ (SE=0.53), $p = .893$ | | | | | |
| SMH ($n = 38$) | | | $\Delta = +0.59 \text{ mm}^3$ (SE=0.50), $p = .241$ | | | | | |

Within-group standardized volume changes from baseline to follow-up were small in both MIG patients and CTRL participants, with confidence intervals including zero in both groups. Overall, these findings suggest that the left anterior-superior hypothalamic subunit remained broadly stable over time, with no evidence of a differential longitudinal change between migraineurs and controls.

The sensitivity analysis restricted to participants scanned with the same coil across acquisitions yielded the same null results (*p* = .873), indicating that the results were not driven by coil changes. The left posterior subunit showed the strongest exploratory signal, with a nominal group-by-time interaction that did not survive FDR correction across the 10 lateralized subunits (*p*_FDR_ = .068). However, within-group standardized changes were small and their confidence intervals included zero, indicating no robust longitudinal change in either MIG patients or CTRL participants. The exploratory comparison of treatment arms showed no evidence of differential longitudinal trajectories between MBSR+ and SMH on the primary outcome.

Overall, these results indicate that the left anterior-superior hypothalamic enlargement observed in MIG patients at baseline did not show evidence of differential normalization over the follow-up period relative to CTRL participants.

## 4. Discussion

The present study examined volumetric changes in hypothalamic subunits in two independent cohorts of patients with episodic migraine (DS1-MIG and DS2-MIG), each compared with its respective CTRL group. To determine whether these structural alterations were specific to migraine, rather than reflecting non-specific adaptations related to pain more broadly or to trigeminal pain specifically, we further examined two clinical control cohorts comprising patients with fibromyalgia (DS-FM) and primary trigeminal neuralgia (DS-TN). In addition, to assess the stability of the observed hypothalamic alterations over time, we examined longitudinal MRI data available for a subsample of DS1-MIG participants.

Three main findings emerged. First, both migraine cohorts showed significant multivariate differences in hypothalamic subunit volume profiles relative to healthy controls, whereas no effects were observed in the fibromyalgia or trigeminal neuralgia cohorts. Second, lateralized follow-up analyses identified a reproducible enlargement of the left anterior-superior hypothalamic subunit across the two migraine datasets. Third, in the longitudinal subsample of the migraine dataset DS1-MIG, this left anterior-superior enlargement showed no evidence of normalization over the follow-up period, despite patients undergoing treatment interventions associated with reduced headache attack frequency. These results remained robust across multiple sensitivity analyses. Specifically, we verified that the residualization procedure used to correct hypothalamic volumes for eTIV adequately accounted for potential age- and sex-related effects. Furthermore, we repeated the main multivariate analyses without outlier exclusion across all datasets, assessed the potential influence of coil type in DS1-MIG in both cross-sectional and longitudinal studies, and examined whether peri-ictal status affected the results in DS2-MIG. Taken together, these findings indicate hypothalamic macrostructural alterations in episodic migraine, while refining current models of hypothalamic involvement by suggesting that these changes are not expressed as a global hypothalamic abnormality, but rather as a region-specific and relatively stable alteration localized to the left anterior-superior hypothalamic subunit. Importantly, this pattern does not appear to reflect non-specific effects of chronic pain or trigeminal pain.

These results are consistent with extensive functional imaging evidence implicating the hypothalamus across the migraine cycle, while also aligning with studies suggesting a more specific involvement of anterior hypothalamic regions. The first functional imaging evidence linking the hypothalamus to migraine came from Denuelle et al. (2007), who reported anterior hypothalamic activation during spontaneous migraine attacks. Subsequent studies in episodic migraine implicated the hypothalamus during the premonitory/preictal phase, including increased hypothalamic perfusion before pain onset (Maniyar et al., 2014) and enhanced functional coupling with the spinal trigeminal nuclei before attacks, followed by altered connectivity with the dorsal pons during the ictal phase (Schulte and May, 2016). Further support for anterior hypothalamic involvement comes from chronic migraine studies, in which stronger anterior hypothalamic activation was reported in chronic migraine compared with episodic migraine and healthy controls during painful trigeminal stimulation, whereas more posterior hypothalamic regions appeared more closely related to the acute headache state (Schulte et al., 2017). In line, an increased connectivity was found between the anterior hypothalamus and the spinal trigeminal nucleus in chronic migraine patients (Lerebours et al., 2019). More importantly, a longitudinal neuroimaging over 30 days in episodic migraine patients demonstrated hypothalamic activation within the 48 hours preceding spontaneous migraine attacks, with activation maxima localized to the lateral anterior hypothalamus (Schulte et al., 2020). This localization appears broadly consistent with the anterior hypothalamic subunit identified in the present study, although with opposite lateralization.

Against this functional background, the interpretation of our findings becomes more complex when considered alongside recent structural investigations using the same hypothalamic segmentation approach. Giardina et al. (2025) found no significant volumetric differences in episodic migraine without aura during strictly interictal periods, while also controlling for menstrual-cycle phase by scanning female participants at mid-cycle. Xiong et al. (2025) reported hypothalamic volume reductions primarily in chronic migraine patients, specifically in the right superior tubular subunit and in inferior hypothalamic regions, including the left inferior tubular and right anterior inferior subunits.

Descriptive analyses suggested a gradual decrease in hypothalamic volumes from controls to episodic and chronic migraine patients within the inferior tubular subunit, although the episodic migraine group did not reach statistical significance, likely due to limited sample size. Notably, in the same study, a Mendelian randomization analysis suggested that smaller volumes of the inferior tubular subregion are linked to increased migraine risk.

These marked discrepancies with our results may reflect both clinical and methodological differences across cohorts. From a clinical perspective, differences in disease burden and timing relative to the migraine cycle may influence the detectability and direction of hypothalamic volumetric alterations. Direct comparisons of disease burden across the reported studies (Giardina et al., 2025; Xiong et al., 2025) and the datasets employed in the present investigations (i.e., DS1-MIG and DS2-MIG) are limited by heterogeneous clinical reporting. Notably, only Giardina et al. (2025) strictly controlled for timing relative to the migraine cycle, excluding patients who experienced an attack within three days before or after the MRI, and for hormonal status, scanning female participants at mid-cycle. From a methodological perspective, it is plausible that the small size of hypothalamic subfields makes volumetric estimates sensitive to MRI acquisition protocol and quality-control procedures. In this context, it is also worth noting that a recent exploratory preprint reported no clear migraine-versus-control volumetric differences, but found that vestibular symptom burden within migraine patients was positively associated with anterior-superior hypothalamic volume, with side-specific analyses suggesting a stronger left-sided effect (Dong et al., 2026). Although preliminary and not peer-reviewed, this finding cautiously converges with our localization to the left anterior-superior subunit and suggests that this territory may be relevant to migraine-related symptom dimensions beyond pain itself.

Although MRI segmentation cannot resolve individual hypothalamic nuclei with histological precision, the anterior-superior hypothalamic subunit is anatomically associated with the paraventricular nucleus (PVN) and the preoptic area (Billot et al., 2020). Given the strong mechanistic evidence implicating the PVN in autonomic-endocrine regulation, oxytocinergic signaling, and trigeminovascular modulation, all highly relevant to migraine pathophysiology, our interpretation here mainly focuses on this nucleus as a central integrative hub for stress-related, metabolic, circadian, and autonomic signals (Iremonger and Power, 2025). Within this broader functional framework, oxytocinergic signaling is particularly relevant, as OT beyond inducing uterine contractions during childbirth and lactation, modulates social behavior, memory, mood, and anxiety (Yao and Kendrick, 2025), and has a prominent role in pain modulation (Juif and Poisbeau, 2013).

PVN and neuropeptide OT seem to play an important role in headache mechanisms (Robert et al., 2013), thus providing a plausible neurobiological interpretation for the anterior-superior hypothalamic enlargement observed in our study. PVN neurons have been shown to project to the superior salivary nucleus and to the caudal spinal trigeminal nucleus (Sp5C) (Robert et al., 2013). Animal studies demonstrated that PVN stimulation inhibits nociceptive trigeminocervical complex neurons responsive to meningeal and periorbital stimulation through oxytocinergic transmission, an effect abolished by local OT receptor antagonism (Condés-Lara et al., 2024). Along these lines, OT has been shown to modulate trigeminocervical complex activity induced by meningeal electrical stimulation, and that its receptors are widely represented in the trigeminal ganglion, a key structure in migraine pathophysiology (Warfvinge et al., 2020). Within this framework, OT administration has been shown to reduce migraine pain (Tzabazis et al., 2017). Notably, a recent randomized, placebo-controlled provocation study investigating acute OT receptor blockade in episodic migraine found that short-acting OT antagonism did not significantly increase migraine-like attacks (Fitzek et al., 2026). These findings suggest that putative acute suppression of OT signaling was not sufficient to provoke migraine-like attacks. However, this remains compatible with a modulatory role of OT within the trigeminovascular system, particularly given the short duration of receptor blockade and the uncertain central availability of atosiban (Fitzek et al., 2026).

The biological interpretation of hypothalamic subunit enlargement remains necessarily cautious, as T1-weighted MRI cannot distinguish among underlying cellular or tissue-level mechanisms. In other contexts, such as elevated body mass index (BMI), larger hypothalamic volume has been discussed as potentially compatible with inflammation-related mechanisms, including gliosis, astrocytic hypertrophy, microglial proliferation, vascular changes, or blood-brain barrier alterations (Brown et al., 2023). Moreover, the consistent left-lateralization of our findings should be interpreted cautiously. Although lateralization of PVN-related oxytocinergic mechanisms in migraine remains unknown, emerging evidence suggests that PVN neuropeptidergic organization may exhibit side-and subregion-specific asymmetries, including asymmetric coordination of vasopressin neuronal populations (Watanabe et al., 2026).

Importantly, the replicated enlargement of the lateralized anterior-superior hypothalamic subunit observed here converges with previous evidence from chronic cluster headache, where an enlargement of the same subunit ipsilateral to the pain was reported (Ferraro et al., 2024). This convergence is notable because, although migraine and cluster headache are distinct primary headache disorders, both show prominent hypothalamic involvement related to attack cyclicity, autonomic-endocrine regulation, and homeostatic modulation (Leone and Proietti Cecchini, 2017; Wei and Goadsby, 2021; May and Burstein, 2019). At the same time, the clinical interpretation differs across disorders: the anterior-superior alteration was observed in chronic cluster headache but not in episodic cluster headache, whereas in our study, it was detectable in episodic migraine.

Perhaps most importantly, the absence of comparable hypothalamic alterations in fibromyalgia and, especially, trigeminal neuralgia suggests that the observed enlargement is unlikely to reflect a generic hypothalamic alteration related to chronic pain or trigeminal pain per se. The trigeminal neuralgia results are particularly informative for interpreting the migraine findings because, although both conditions involve the trigeminal system, they differ substantially in their broader pathophysiological architecture. Migraine is characterized by cyclic fluctuations, premonitory symptoms, and autonomic and neuroendocrine manifestations, all of which strongly implicate hypothalamic circuits (May and Burstein, 2019). In contrast, trigeminal neuralgia is primarily characterized by recurrent severe paroxysmal pain occurring within the trigeminal nerve distribution, reflecting involvement of the trigeminal somatosensory system (Bendtsen et al., 2020). Thus, the absence of similar hypothalamic changes in trigeminal neuralgia supports the interpretation that our findings are more closely related to migraine-specific hypothalamic mechanisms than to trigeminal nociception alone.

Beyond the replicated left anterior-superior effect, the DS2-MIG cohort also showed additional enlargements of the right anterior-inferior and right tubular-inferior subunits. Because these additional effects did not replicate in DS1-MIG, they should be interpreted cautiously, in particular for the anterior-inferior subunit, which showed the lowest reproducibility among all the subunits in the original validation study (Billot et al., 2020). As discussed above in relation to discrepancies with previous studies, between-cohort differences in clinical characteristics (such as migraine frequency, attack duration, and disease duration), timing relative to the migraine cycle, and MRI acquisition parameters may increase variability in hypothalamic subfield estimates and may partly explain why these additional effects were observed only in DS2-MIG. Thus, unlike the left anterior-superior enlargement, which was observed across both migraine cohorts, the DS2-MIG-specific anterior-inferior and tubular-inferior effects should be considered exploratory until replicated in larger independent samples.

The absence of longitudinal normalization observed in the DS1-MIG dataset raises the question of whether the left anterior-superior hypothalamic enlargement should be interpreted as a consequence of migraine or as a predisposing factor. One possibility is that recurrent migraine attacks, or migraine-related homeostatic and autonomic dysregulation more broadly, induce long-lasting structural adaptations in this region. Alternatively, this alteration may precede migraine onset and reflect a vulnerability marker, whereby individuals with specific hypothalamic macrostructural features are more susceptible to developing migraine. Because the available longitudinal data were acquired after disease onset and did not include pre-morbid imaging, the present study cannot distinguish between these scenarios. Nevertheless, the persistence of the effect over follow-up in the DS1-MIG dataset, despite reduced headache frequency, suggests that the alteration is unlikely to represent a purely state-dependent marker of current attack burden. Rather, it may reflect a relatively stable migraine-related feature.

Particularly relevant to the present findings, a recent study in chronic insomnia disorder (Luo et al., 2024b) identified enlargement of the left anterior-superior hypothalamic subunit using the same automated hypothalamic segmentation framework. The authors further reported associations between hypothalamic hypertrophy, symptom severity, and circulating CRH levels, highlighting a potential role of HPA-axis dysfunction. Given the well-established links between migraine, sleep disturbances, stress responsivity, and neuroendocrine regulation, these observations are consistent with the hypothesis that enlargement of the anterior-superior hypothalamic territory may reflect alterations in stress–homeostatic systems.

### 4.1. Limitations

Several limitations should be acknowledged. First, we were not able to assess the potential effects of medication use and headache burden due to the limited availability and heterogeneity of clinical information across the migraine datasets. Regarding medication, the original DS1-MIG study (Seminowicz et al., 2020) reported that only a minority of patients were using preventive migraine treatments at baseline, and individuals using opioid medications were excluded. In addition, in our cohort (DS2-MIG), participants were not receiving prophylactic medications and had not taken acute migraine treatments in the 24 hours preceding MRI acquisition. Altogether, these observations reduce the likelihood that the observed hypothalamic alterations were primarily driven by pharmacological effects. Regarding headache burden, although it could not be directly modelled across datasets, the presence of left anterior-superior enlargement in the same direction in both migraine cohorts (DS1-MIG and DS2-MIG), despite likely differences in clinical burden, suggests that this alteration may reflect a relatively stable trait-like feature of episodic migraine rather than a state-dependent marker of current headache burden. This interpretation is further supported by the longitudinal DS1-MIG data: the enlargement showed no detectable tendency toward normalization despite reductions in mean headache days, from approximately 7.8 to 4.6 days per 28-day period in the MBSR+ group and from 7.7 to 6.0 days in the SMH group between baseline and week 20. In addition, no detectable differences in longitudinal volumetric change were observed between the two treatment groups, despite their different post-treatment headache burden.

Second, the timing of MRI acquisition relative to the migraine cycle was not available for the DS1-MIG dataset. This represents a relevant limitation, as hypothalamic activity and connectivity are known to fluctuate across the migraine cycle. Importantly, in the DS2-MIG dataset, no migraine participant was scanned during an ongoing attack, and sensitivity analyses (excluding patients with an attack occurring 3 days before the MRI) supported the consistency of the reported findings. However, headache occurrence after the MRI session was not systematically assessed; therefore, we cannot exclude that some participants classified as non-ictal may have been scanned during a peri-ictal or premonitory phase.

Third, BMI and other metabolic variables were not available and were therefore not included as covariates. This represents a relevant limitation, because the hypothalamus is strongly involved in energy balance, appetite regulation, endocrine function, and metabolic homeostasis. Accordingly, we cannot exclude the possibility that unmeasured metabolic factors may have contributed to inter-individual variability in hypothalamic subunit volumes.

### 4.2. Conclusion

In conclusion, our findings indicate that episodic migraine is associated with a reproducible, subunit-specific enlargement of the left anterior-superior hypothalamus. The absence of comparable effects in fibromyalgia and trigeminal neuralgia suggests that this alteration is not simply explained by chronic pain or trigeminal involvement more broadly. Rather, it may reflect a migraine-relevant macrostructural alteration of an anterior hypothalamic territory involved in homeostatic, autonomic, endocrine, and trigeminovascular regulation. Its replication across independent migraine cohorts and persistence over longitudinal follow-up support its interpretation as a relatively stable structural marker of hypothalamic involvement in episodic migraine.

**Figure 1:**
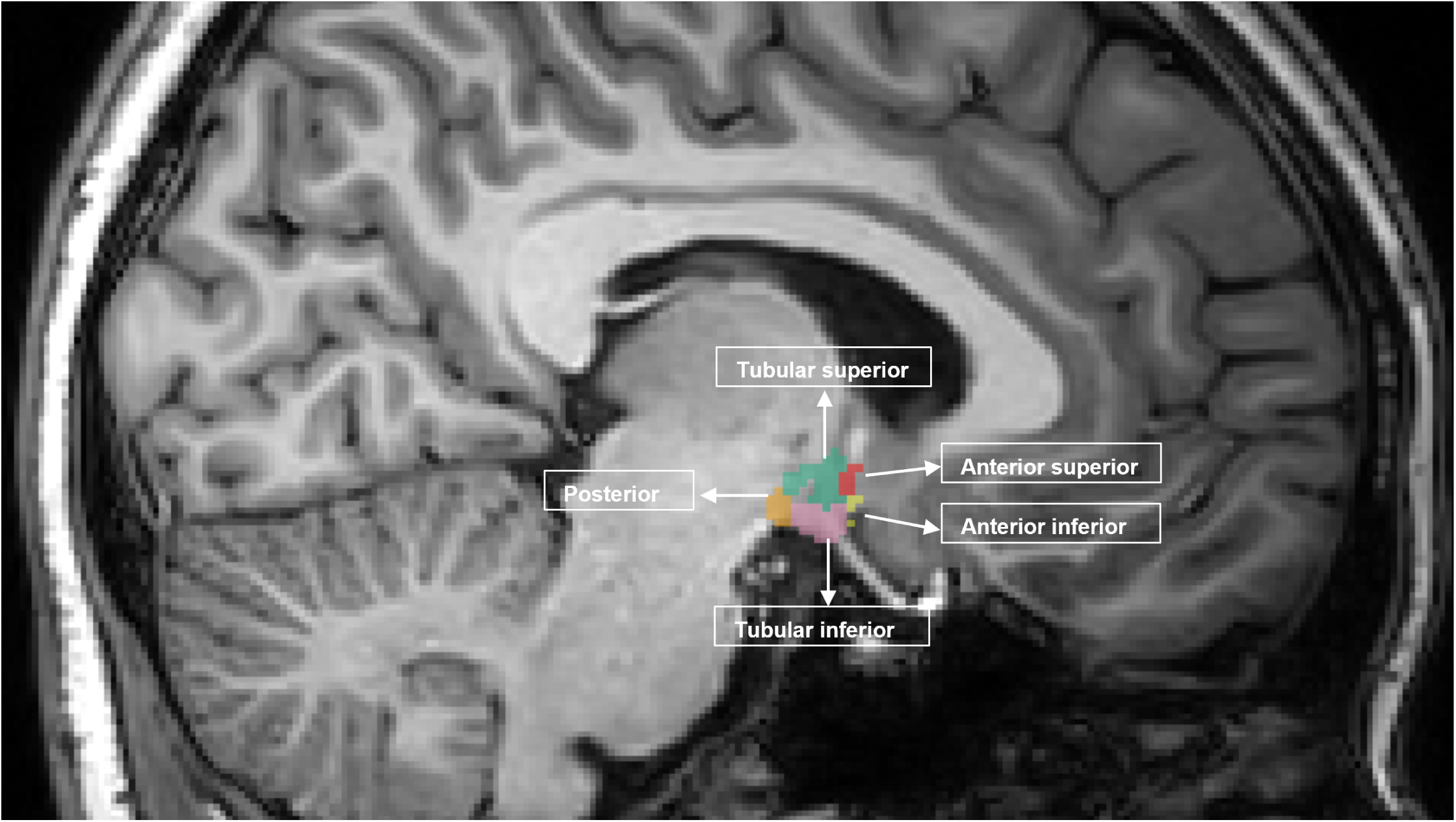
T1-weighted MRI sagittal section illustrating the segmentation and anatomical localization of the hypothalamic subunits of a representative subject.

**Figure 2:**
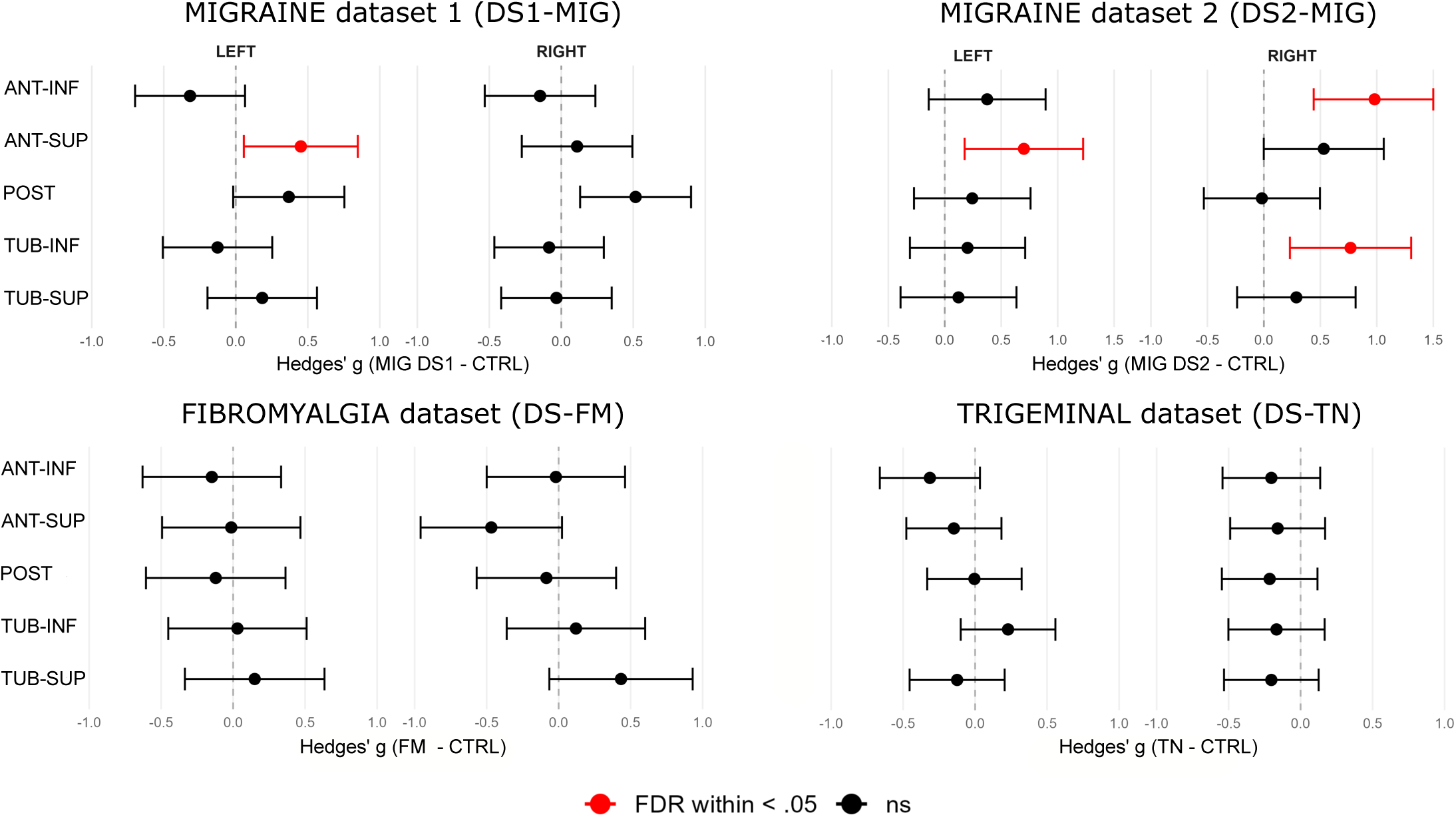
Effect sizes for group differences in eTIV-adjusted hypothalamic subunit volumes across four cohorts. Forest plots show Hedges’ *g* (patient minus control) with 95% CI for each subunit, separately for the left and right hemispheres, in the migraine (DS1-MIG-base and DS2-MIG), fibromyalgia (DS-FM), and trigeminal neuralgia (DS-TN) cohorts. Red dots indicate FDR-significant effects (*p*_FDR_ < .05); *g* > 0 reflects larger volumes in the patient group. Significant enlargements were restricted to the migraine cohorts: left anterior-superior in the DS1-MIG dataset, and left anterior-superior, right anterior-inferior, and right tubular-inferior in the DS2-MIG dataset. No significant differences were observed in the DS-FM or DS-TN datasets.

**Figure 3:**
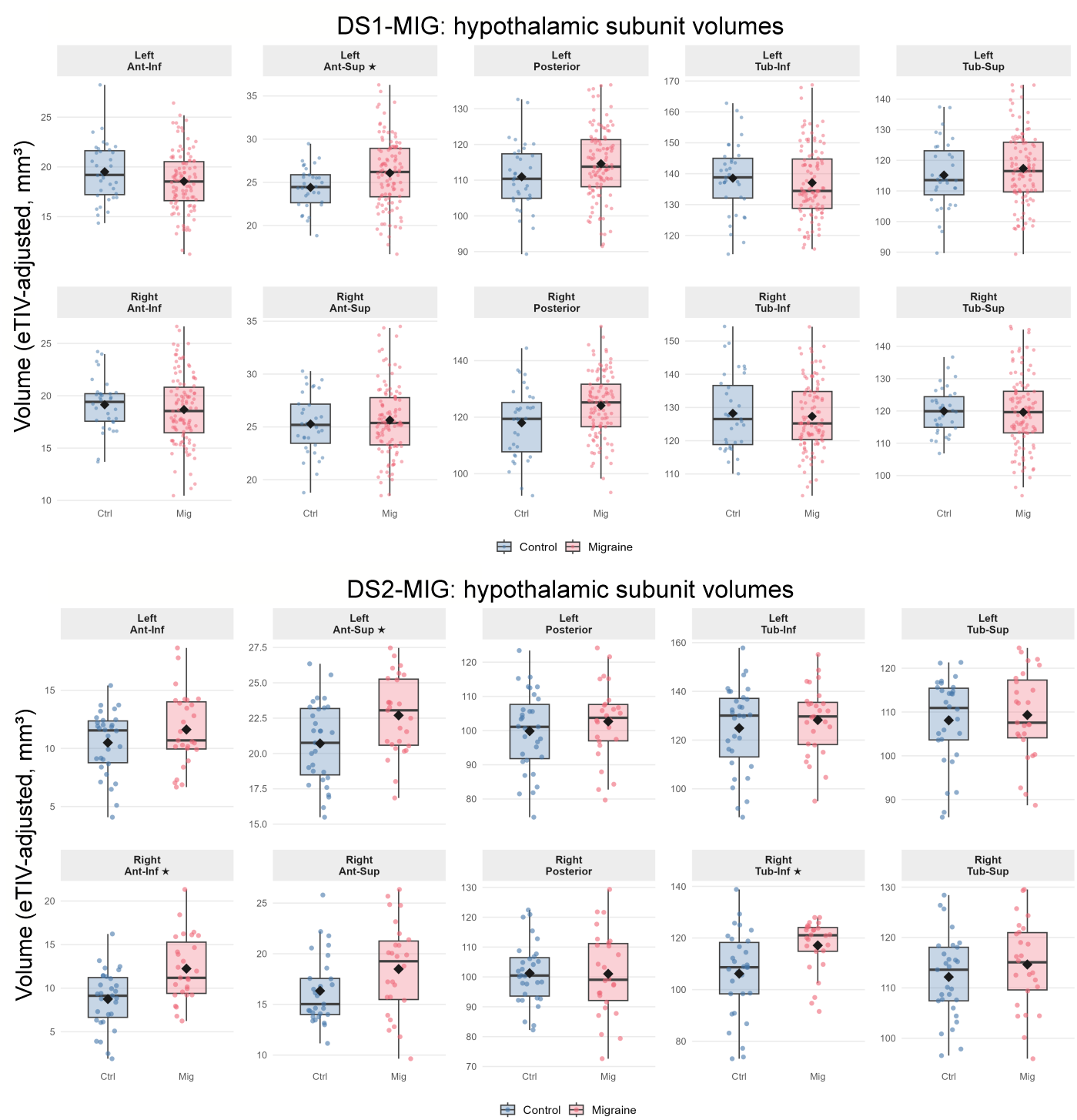
Boxplots show the distribution of eTIV-residualized volumes (mm^3^) for Control (blue) and Migraine (pink) groups in the DS1-MIG dataset and DS2-MIG dataset, separately for left and right hemispheres. Diamonds indicate group means; dots represent individual observations. Subunits reaching FDR significance are marked with ⋆; In DS1-MIG, the left anterior-superior subunit was significantly larger in migraine patients. In DS2-MIG, significant enlargements were observed in the right anterior-inferior and right tubular-inferior subunits, with a significant left anterior-superior effect.

**Figure 4:**
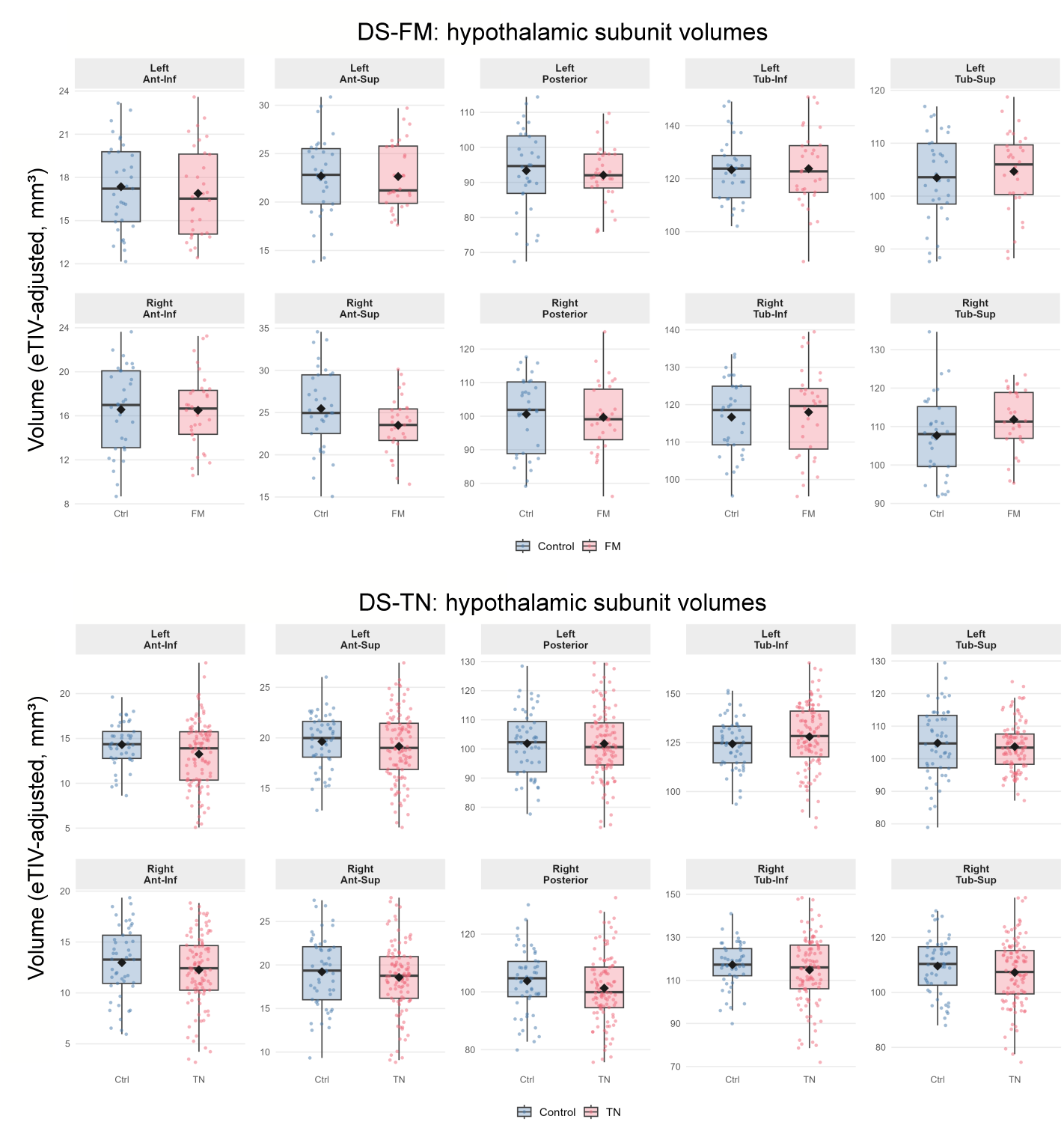
Boxplots show the distribution of eTIV-residualized volumes (mm^3^) for Control (blue) and Patients (pink) groups in the DS-FM dataset and DS-TN dataset, separately for left and right hemispheres. Diamonds indicate group means; dots represent individual observations. No subunit reached statistical significance in either cohort after FDR correction.

**Figure 5:**
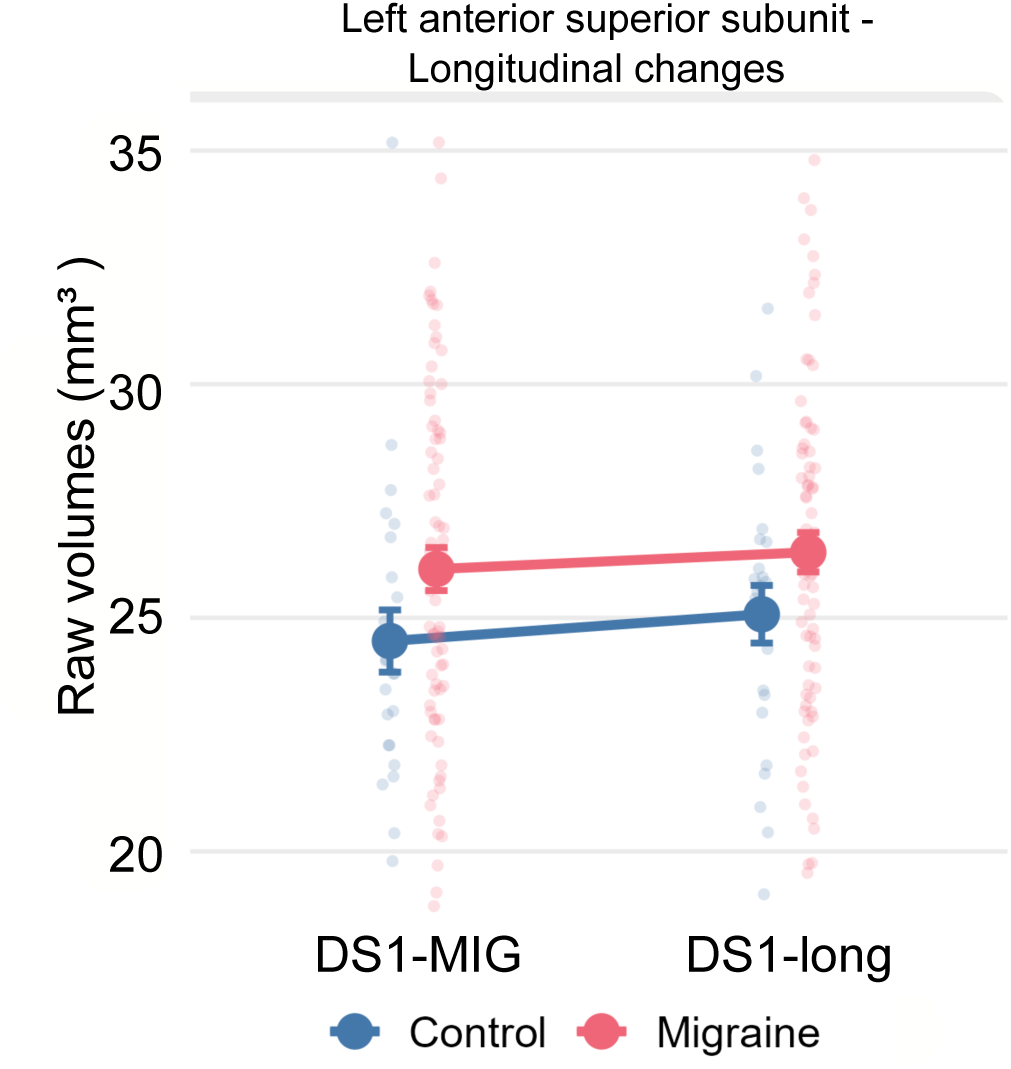
Longitudinal changes in left anterior superior hypothalamic subunit volume. Raw volumes (mm^3^) are shown at baseline and follow-up for the Migraine (pink) and Control (blue) groups. Large symbols represent group means ± standard error; small dots represent individual observations. The between-group difference observed at baseline (Hedges’ *g* = +0.45, 95% CI [0.06, 0.85]) was not significantly modified over time (*p*_interaction_ = .754).

## Acknowledgements

This study used openly available datasets obtained from the OpenNeuro repository. We gratefully acknowledge the original investigators and participants who contributed to these datasets and made them available to the scientific community through open data sharing initiatives.

## Data availability

Data are available upon request.

## Supplementary Material

**Table SM1:** MRI acquisition parameters for the structural datasets included in the study. All datasets included 3D T1-weighted structural images. Because DS1-MIG included two scanner/head-coil configurations, the potential influence of this acquisition factor was explicitly assessed through sensitivity analyses in both the DS1-MIG cross-sectional and longitudinal analyses.

| Dataset | Scanner | Head coil / coil distribution | TR (ms) | TE (ms) | Flip angle | FOV / Voxel size |
| --- | --- | --- | --- | --- | --- | --- |
| DS1-MIG | 3.0 T Siemens Trio / Prisma | DS1-MIG-base: 32-channel, 111 MIG and 21 CTRL; 64-channel, 14 CTRL. DS-MIG-long: 12 participants were scanned with a different coil relative to baseline. | 2300 | 2.98 | 9° | 256 × 256 × 170 mm<br>1 × 1 × 1 mm |
| DS2-MIG | 3.0 T Philips Ingenia | 32-channel | 8.19 | 3.78 | 8° | 240 × 240 × 180 mm<br>1 × 1 × 1 mm |
| DS-FM | 3.0 T Philips Ingenia | 32-channel | 7.0 | 3.5 | 8° | 240 × 240 mm<br>1 × 1 × 1 mm |
| DS-TN | 3.0 T Philips Ingenia | 16-channel | 6.57 | 2.95 | 8° | 256 × 256 mm<br>1 × 1 × 1 mm |

**Table SM2:** Sensitivity analysis: effect of eTIV residualization on sex- and age-related variance in hypothalamic subunit volumes (controls only). Sex analysis: Welch *t*-test comparing males vs. females on raw and eTIV-adjusted bilateral volumes. Age analysis (DS2, *n* = 31; DS-FM, *n* = 33): Spearman *ρ* between bilateral volumes and age, before and after eTIV adjustment.

| Sex effects before and after eTIV adjustment |  |  |  |  |
| --- | --- | --- | --- | --- |
| Subunit | DS1-MIG-base (CTRL: $n_M = 4$, $n_F = 30$) | | DS2-MIG (CTRL: $n_M = 17$, $n_F = 14$) | |
| | $p_{\text{raw}}$ | $p_{\text{adj}}$ | $p_{\text{raw}}$ | $p_{\text{adj}}$ |
| Anterior-inferior | .091 | .145 | .057 | .445 |
| Anterior-superior | .172 | .270 | .488 | .548 |
| Posterior | .122 | .699 | .086 | .751 |
| Tubular inferior | .089 | .600 | .073 | .242 |
| Tubular superior | .198 | .644 | .070 | .925 |

| Age effects before and after eTIV adjustment |  |  |  |  |
| --- | --- | --- | --- | --- |
| Subunit | DS2-MIG (CTRL: $n = 31$) | | DS-FM (CTRL: $n = 33$) | |
| | $\rho_{\text{raw}} (p)$ | $\rho_{\text{adj}} (p)$ | $\rho_{\text{raw}} (p)$ | $\rho_{\text{adj}} (p)$ |
| Anterior-inferior | + .018 (.923) | − .039 (.833) | − .067 (.713) | − .050 (.781) |
| Anterior-superior | + .061 (.743) | + .056 (.763) | + .109 (.548) | + .200 (.266) |
| Posterior | − .210 (.256) | − .110 (.558) | + .138 (.445) | + .232 (.193) |
| Tubular inferior | − .373 (.039) | − .334 (.067) | − .133 (.461) | − .113 (.531) |
| Tubular superior | − .263 (.153) | − .175 (.346) | + .164 (.361) | + .268 (.132) |

**Table SM3:** Dataset 1 Coil 1-only MANOVA sensitivity analysis. The five bilateral ICV-adjusted hypothalamic sub-units were entered as joint dependent variables, with group (Migraine vs. Control) as the between-subjects factor. Pillai’s trace is reported as the primary multivariate test statistic. The primary Coil 1 model used the same hybrid outlier-handling strategy as the main analysis, with listwise deletion across the five dependent variables. The sensitivity model retained single-unit values while excluding multi-unit outlier subjects.

| **Model** | **n** | **Pillai** | **F (df)** | **p** |
| --- | --- | --- | --- | --- |
| Primary Coil 1, hybrid + listwise | 118 | 0.126 | 3.21 (5, 112) | .010 |

**Table SM4:** DS2-MIG dataset peri-ictal sensitivity analysis of the bilateral omnibus MANOVA. The five bilateral ICV-adjusted hypothalamic sub-units were entered as joint dependent variables, with group (Migraine vs. Control) as the between-subjects factor. Pillai’s trace is reported as the primary multivariate test statistic. The reference model includes all migraineurs scanned within 3 days from their last attack. MANCOVA models include biological sex as a categorical covariate. Abbreviations: CTRL = healthy controls; MIG = migraine.

| **Cut-off** | **Model / term** | **n (CTRL/MIG)** | **Pillai** | **F (df)** | **p** |
| --- | --- | --- | --- | --- | --- |
| $\geq 3$ days | MANOVA, group | 43 (28/15) | 0.356 | 4.09 (5, 37) | .005 |
|  | MANCOVA, group \| sex | 43 (28/15) | 0.371 | 4.25 (5, 36) | .004 |
|  | MANCOVA, sex \| group | 43 (28/15) | 0.033 | 0.25 (5, 36) | .940 |

